# Genomic alterations in the YAP/TAZ pathway are associated with stem cell-like castration-resistant prostate cancer

**DOI:** 10.1101/2025.07.01.25330467

**Authors:** Marjorie L. Roskes, Alexander Martinez-Fundichely, Sandra Cohen, Metin Balaban, Chen Khuan Wong, Weiling Li, Tonatiuh A. Gonzalez, Anisha B. Tehim, Hao Xu, Shahd ElNaggar, Matthew Myers, Rohan Bareja, Princesca Dorsaint, Kathryn Gorski, Muhammad Asad, Majd Al Assaad, Brian D. Robinson, Michael Sigouros, Ethan Barnett, Jyothi Manohar, Scott Tagawa, David Nanus, Ana Molina, Jones T. Nauseef, Cora N. Sternberg, Juan Miguel Mosquera, Howard I. Scher, Andrea Sboner, Benjamin J. Raphael, Yu Chen, Ekta Khurana

## Abstract

Castration-resistant prostate cancer (CRPC) is an aggressive disease exhibiting multiple epigenomic subtypes: androgen receptor-dependent CRPC-AR, and lineage plastic subtypes CRPC-SCL (stem cell-like), CRPC-WNT (Wnt-dependent), and CRPC-NE (neuroendocrine). By transcriptomic profiling of tissue, and whole-genome sequencing (WGS) of tissue and cell-free DNA (cfDNA) from 500 samples, we relate genomic variants with epigenomic state. We find lineage plasticity is associated with higher epigenomic and genomic heterogeneity. Samples with CRPC-SCL show higher chromosomal instability. We find DNA alterations, particularly chromosomal rearrangements, in the YAP/TAZ pathway associated with CRPC-SCL. For example, complex rearrangements on chromosome 4, which are supported by patient-matched 3D genome architecture data, decrease promoter interactions of *MOB1B*, a YAP/TAZ pathway inhibitor, with its enhancers. Together, the genomic variants in the pathway can predict CRPC-SCL with 79% accuracy. We show the utility of cfDNA WGS for joint inference of epigenomic state and genomic variants, which can guide patient stratification for clinical decisions.

**Significance:** This study reveals genomic variants associated with the presence of lineage-plastic CRPC stem cell-like state. We leverage the utility of minimally invasive cfDNA sequencing to obtain genomic and epigenomic insights about CRPC heterogeneity, which have implications for patient stratification for treatment decisions.

## Introduction

In the treatment-naïve stage, the growth of prostate cancer is largely dependent on the androgen receptor (AR) pathway. As such, the first line of treatment for advanced prostate cancer is androgen deprivation therapy (ADT). Tumors often develop resistance to ADT and are termed castration-resistant prostate cancer (CRPC). Thus, often next generation AR signaling inhibitors (ARSIs)^1–4^ are used in combination with ADT as a first-line therapy. ARSIs work to inhibit AR signaling by different mechanisms. For example, abiraterone inhibits steroidogenesis of adrenal androgens, while enzalutamide and apalutamide inhibit translocation of the AR dimer to the nucleus thereby blocking DNA and cofactor binding^5^. Selective pressure can result in tumors that acquire resistance to ARSIs, either by way of genomic alterations of AR such as amplification of the AR gene or alternative splicing of AR mRNA, or by circumventing dependence on AR altogether^5^. While some of these lineage plastic, AR-independent tumors transdifferentiate into a state with neuroendocrine histology, others remain in the non-AR, non-neuroendocrine state, often referred to as “double-negative” prostate cancer, retaining their adenocarcinoma histology^6,7^.

By using chromatin and transcriptomic profiling, we identified four subtypes of CRPC: CRPC-AR which depends on the AR pathway, CRPC-SCL which overexpresses stem cell-like markers, CRPC-WNT which is dependent on the WNT signaling pathway, and CRPC-NE which constitutes the neuroendocrine tumors expressing neuroendocrine gene markers^8^. CRPC-WNT and CRPC-SCL tumors also exhibit enrichment of fibroblast growth factor receptor (FGFR) signaling, consistent with previous studies of double-negative tumors^6^. Moreover, CRPC-SCL tumors also exhibit overexpression of Janus kinase/signal transducer and activator of transcription (JAK/STAT) signaling, which has been shown to drive lineage plasticity in CRPC^6,9^. Here, we refer to CRPC-SCL, CRPC-WNT, and CRPC-NE as lineage plastic subtypes since they show divergence from the CRPC-AR state.

The identification of these four subtypes in patient tumors is critical since it can offer therapeutic opportunities^8^. While recent efforts have found evidence of intra-patient CRPC subtype heterogeneity, they were limited by small patient cohorts and did not examine the extent of heterogeneity in the context of all these four subtypes^10,11^. The CRPC-SCL subtype in particular has not been characterized well by previous studies. While genomic events have been associated with CRPC-AR (alterations of the AR gene and enhancer), CRPC-NE (loss of RB1), and CRPC-WNT (alterations in the Wnt signaling pathway), no such events have been described for CRPC-SCL^8,12–17^.

In this study, we seek to fill these gaps via a combined analysis of genomic events and the four epigenomic subtypes, defined by their gene expression and chromatin accessibility profiles. To do so, we annotate the samples with epigenomic subtypes by quantifying the fractional contribution of each using tissue RNA-seq and/or nucleosomal profiling using cell-free DNA (cfDNA) whole-genome sequencing (WGS). We then relate the genomic events with the epigenomic subtypes by analyzing matched WGS and RNA-seq in the tissue cohort, as well as WGS in the cfDNA cohort.

We find novel genomic alterations at the YAP/TAZ pathway genes that are associated with CRPC-SCL. Because genomic alterations can be difficult to interpret in the absence of other data types, we used matched Hi-C, which reveals three-dimensional (3D) genome architecture, to validate their presence and infer their functional impact^18^. Additionally, we find associations of epigenomic subtypes with genomic instability and heterogeneity, as computed from copy number alterations (CNAs) and structural variations (SVs).

## Results

### Overview of patient samples used in the study

To perform a combined analysis of genomic events and epigenomic subtypes, we annotate the samples by quantifying the fractional contribution of each CRPC subtype. This is done by using deconvolution approaches on tissue RNA-seq (n=455) and nucleosome profiling from cfDNA WGS (n=68)^8,10,14,15,19,20^ (fig. 1A, 2A,B). We then relate the genomic events from WGS with the epigenomic subtypes. This is done by using the patient samples for whom both RNA-seq and WGS was performed on tissue (n=105), as well as the cfDNA whole-genomes^14,21^. Since both genomic variants and epigenomic subtypes are inferred from the same WGS data, the cfDNA whole-genomes in particular offer a unique opportunity to examine their relationship at the same tumoral state and timepoint^15^. Moreover, the use of cfDNA whole-genomes offers the opportunity to monitor the progression of tumors into different epigenomic subtypes with the accompanying genomic changes. Additionally, since prostate cancer genomic alterations are dominated by large SVs^22–25^, the use of deep WGS is important to capture the whole spectrum of genomic events as they may relate to chromatin-based subtypes. Due to the limited availability of deep cfDNA WGS of lineage plastic subtypes (n=2 among n=35 patients^15^), we sequenced additional patient samples (n=20) at 40X coverage from Weill Cornell Medicine (WCM) and Memorial Sloan Kettering (MSK) (fig. 1B). For 7 of these 20 samples, we also had RNA-seq from tissue obtained at the same timepoint as plasma, allowing comparison of molecular subtypes inferred from tissue RNA-seq versus cfDNA WGS. cfDNA whole-genomes from an additional control cohort of healthy participants (n=14) were sequenced to account for the signal from peripheral blood mononuclear cells (PBMCs) in estimating tumoral subtypes from cfDNA (table S1).

**Fig. 1:**
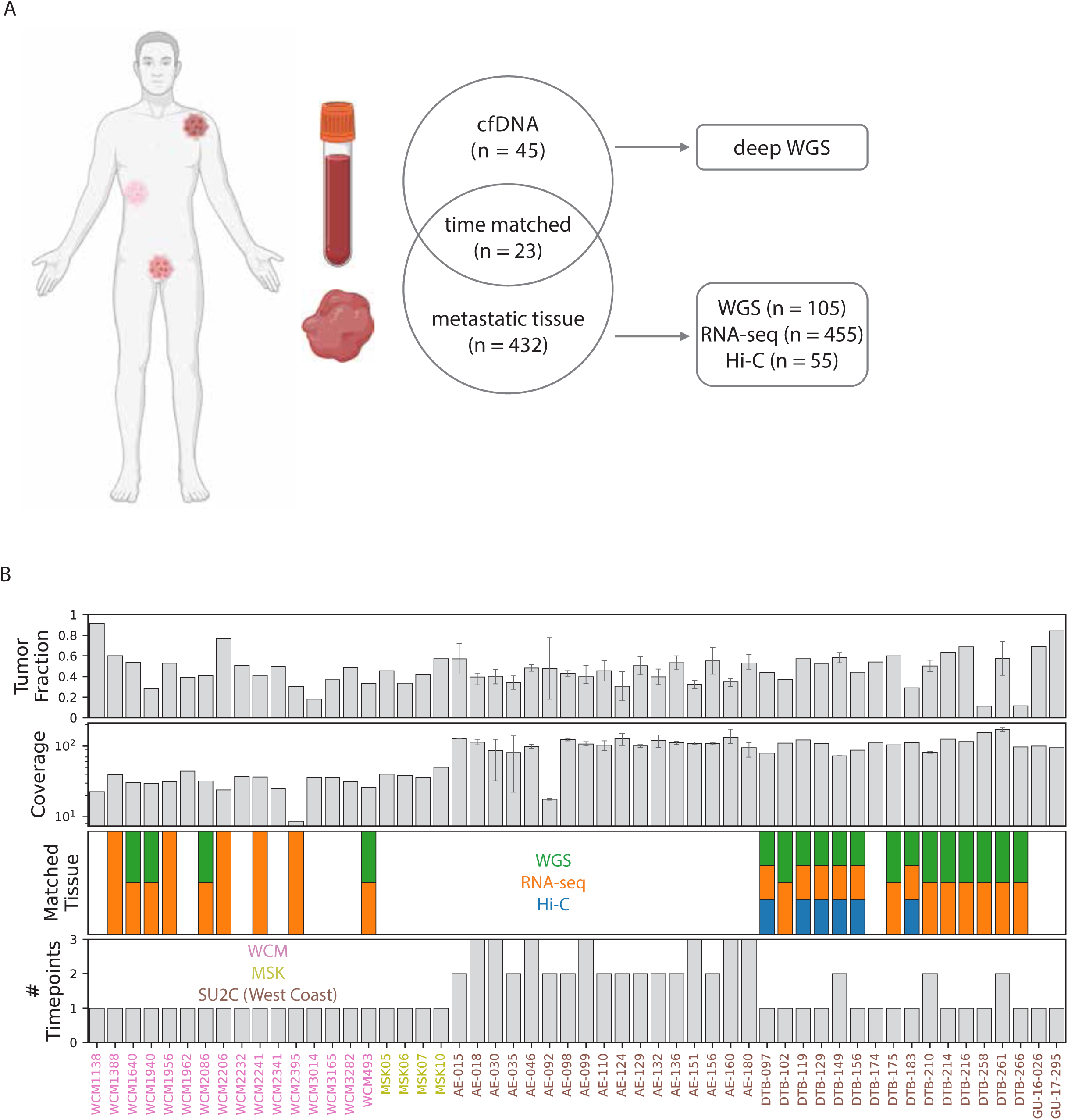
Cohort overview. (A) Number of WGS cfDNA samples from plasma and WGS, RNA-seq, and Hi-C samples from tissue. Some patients have >1 sample. (B) per-sample overview of data availability for cfDNA samples.

### Inter- and intra- Sample CRPC subtype heterogeneity in different histological classes

Due to the concordance of the four molecular subtypes identified from chromatin (ATAC-seq) and transcriptomic (RNA-seq) profiling^8^, we used their signature genes and accessible chromatin sites to infer their presence in patient samples from RNA-seq and cfDNA WGS, respectively. Signature expression genes for each subtype were defined as those showing subtype-specific expression in cell lines/organoids^8^ (tables S2, S3). By comparing expression profiles of subtype-specific signature genes in patient samples to that in cell lines/organoids, we can estimate the fractional contribution of each CRPC subtype (fig. 2A, S1A, S2A) (table S4)^20^. For cfDNA, we leverage the fact that it is enriched for nucleosome-bound regions in tumor tissue^26^. The aggregate coverage profile across cfDNA samples from patient-derived xenografts (PDXs) was computed for subtype-specific accessible chromatin sites. To account for the signal from PBMCs inherent in cfDNA, we generated a panel of no-cancer controls (n=14) (table S5). By comparing coverage profiles in subtype-specific signature sites in patient samples to that of the PDX-derived signature, we can estimate the fractional contribution of each CRPC subtype (fig. 2B, S1B) (tables S6, S7)^10^. The strength of these deconvolution approaches is that they allow assignment of a single sample to a mixture of multiple subtypes.

**Fig. 2:**
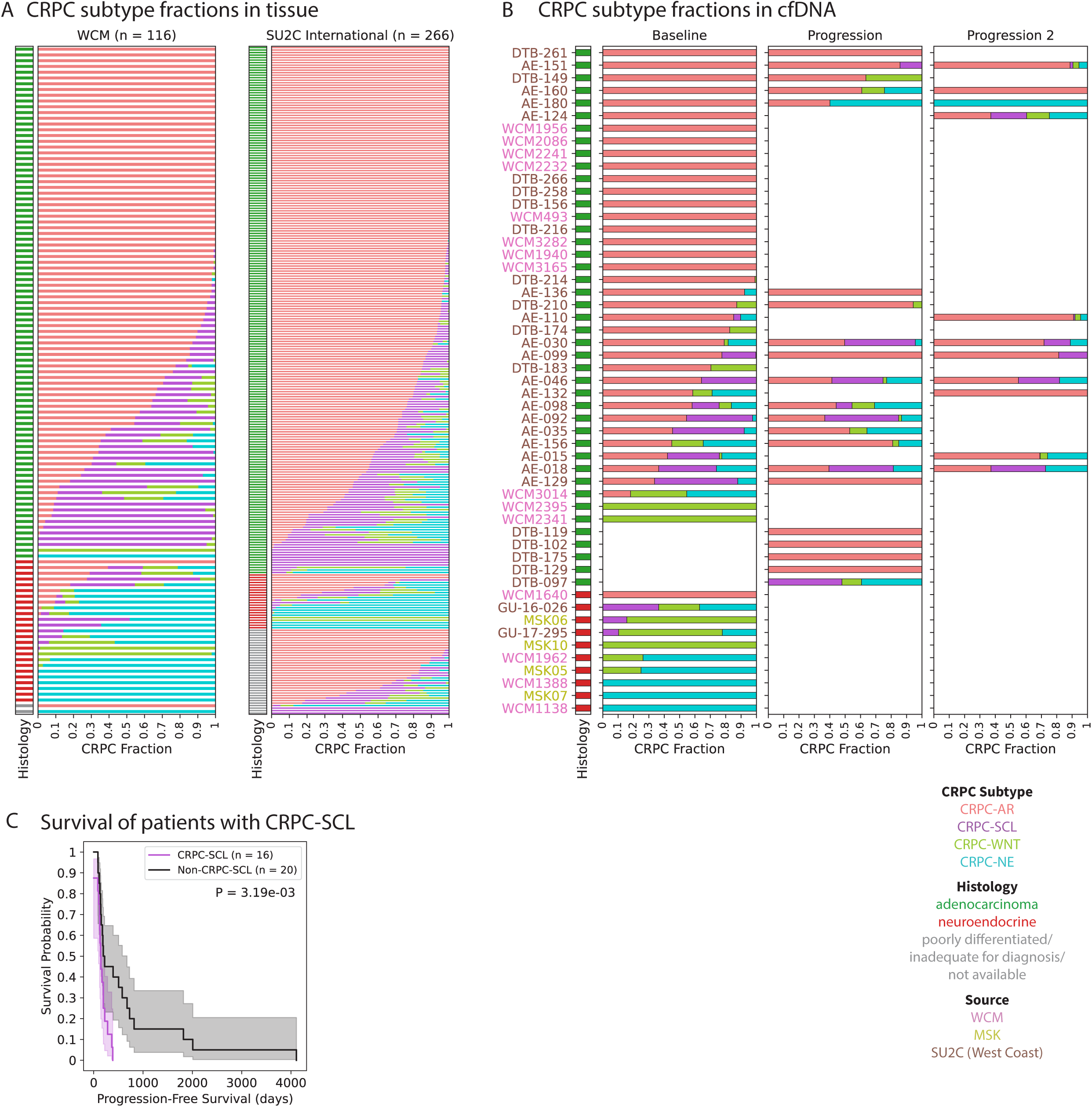
CRPC subtype components in cfDNA and tissue. (A-B) CRPC fractions from deconvolution in tissue (A) and cfDNA (B) cohorts. (C) Kaplan Meier curve showing shorter progression free survival in patients with CRPC-SCL.

To validate the assignment of samples to different subtypes, we checked the expression of the top, well-studied subtype-specific marker genes and pathways (table S8)^8^. As expected, we find that *AR* is upregulated in CRPC-AR samples^27^ (fig. S3A). Interestingly, we find that *TP53* and *PTEN* are downregulated in lineage plastic samples, the mutational loss of which has been reported to be associated with lineage plasticity and aggressive disease in CRPC^8,17,28^ (fig. S3B). Similarly, *RB1* is downregulated while *ASCL1*, *NEUROD1*, *SYP*, and genes in the NEPC pathway are upregulated in CRPC-NE^8,29^ (fig S3C). CRPC-WNT samples show overexpression of genes in the WNT pathway^27^ (fig. S3D), while CRPC-SCL samples show upregulation of many of the YAP/TAZ pathway target genes (fig. S3E)^8^. Furthermore, we find that CRPC-SCL patients show shorter survival on ARSIs than those without CRPC-SCL, consistent with their dependence on transcription factors (TFs) other than AR (fig. 2C)^8^. Furthermore, to confirm that non-tumor cells do not influence the CRPC subtype fraction estimation, we used single-cell RNA-seq (scRNA-seq) data from CRPC patients that allows annotation of each cell as tumoral or non-tumoral^30^. To compare with our approach, we created pseudo-bulk RNA-seq from only tumor cells and all cells. We then computed the fractional components estimated in pseudo-bulk from all cells versus that from tumor cells only and find high concordance, even in samples with tumor fraction as low as 0.13 (fig. 3A, B). This shows that the signature genes and the deconvolution approach used in our study allows accurate assessment of the subtypes from bulk RNA-seq.

**Fig. 3:**
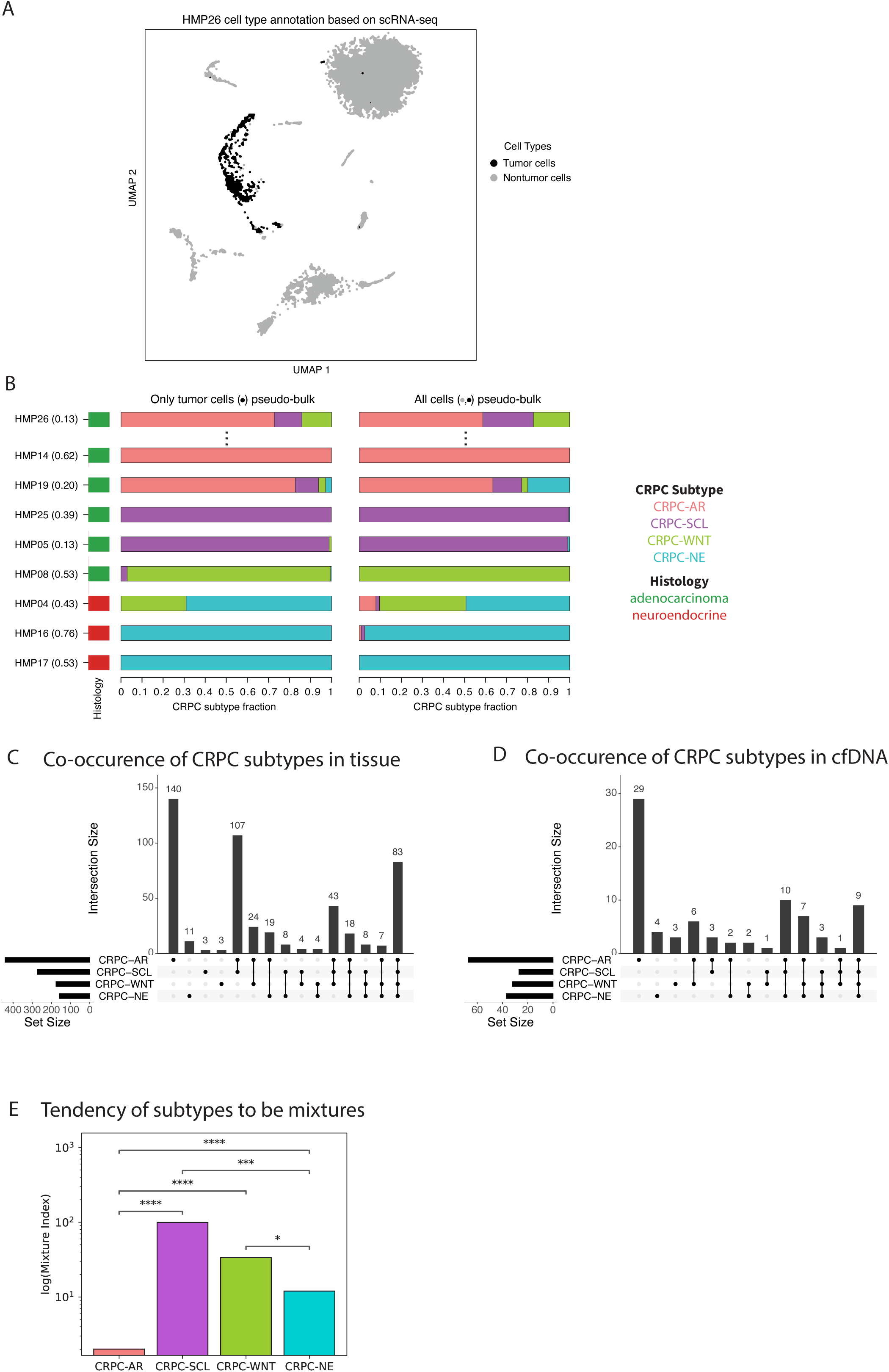
Heterogeneity of CRPC subtypes. (A) Cell type annotation based on scRNA-seq in example sample. (B) CRPC fractions from deconvolution in tumor cells pseudobulk and all cells pseudobulk validates non-tumor cells do not impact subtype inference. (C-D) Upset plots showing number of samples with each CRPC subtype combination in tissue (C) and cfDNA (D). (E) ratio of number of samples with subtype mixtures to pure samples across subtypes.

Although samples are generally classified as adenocarcinoma, neuroendocrine, or mixed based on histology clinically, we find that molecularly they are, in fact, quite heterogeneous^31^. While most of the adenocarcinoma samples (96% for tissue and 94% for cfDNA) showed evidence of CRPC-AR as expected, a large majority show some fraction of other molecular subtypes (76% for tissue and 60% for cfDNA). These include 33% with CRPC-SCL, 28% with CRPC-WNT, and 28% with CRPC-NE (fig. S2B, C). Similarly, while most neuroendocrine samples show evidence of CRPC-NE (89% for tissue and 70% for cfDNA), many of them also show presence of other subtypes. CRPC-SCL is the most common molecular subtype in adenocarcinoma samples after CRPC-AR, and CRPC-WNT is the most common subtype in neuroendocrine samples after CRPC-NE (fig. S2B, C).

We find that the samples with the presence of any of the lineage plastic subtypes tend to be more heterogeneous: these samples most often have co-occurrence of multiple CRPC subtypes. This phenomenon is especially true for the CRPC-SCL subtype, which co-occurs with other subtypes in 99% of cases (fig. 3C, D). We compared the ratio of numbers of samples with mixed subtypes to single subtype and call this the subtype mixture index. We find that CRPC-SCL, CRPC-WNT, and CRPC-NE have significantly higher mixture index values than CRPC-AR samples, which exhibit single subtype in 33% of the cases (fig. 3E). Although CRPC-NE has significantly higher subtype mixture index than CRPC-AR, we find that the samples contributing to the higher index are from the adenocarcinoma histology. In other words, when CRPC-NE exists as the only subtype at the molecular level, it has neuroendocrine histology, while its existence in adenocarcinoma samples is in the presence of other subtypes, mostly CRPC-AR (Fisher’s Exact Odds Ratio = 18.85, P = 1.21e-10) (fig. S2D). This trend is as expected and shows that histology-based classification may miss the beginning of lineage plasticity, which is captured by our molecular annotation. Thus, molecular annotations, when used together with histological classifications, can have clinical relevance by capturing the transition to lineage plasticity, and beginning of treatment resistance.

### Enrichment of subtypes at different metastatic sites

Formation of metastases at distant sites is a key feature of cancer and cancer-related death^32^. In order for a metastasis to form, a tumor cell has to invade the blood stream, survive intense combat with the immune system, exit the bloodstream, and colonize at a distant site^33^. Different tumor cells are more prone to seeding in specific distant sites, a phenomenon known as organ tropism, which is largely influenced by the compatibility of the site-specific microenvironment to the tumor cell^33^. Moreover, the tendencies of tumor cells to metastasize to specific sites can be subtype-specific within a given cancer type^34^. While bone is the most common site of prostate cancer metastasis, other sites are also frequently found. In light of this, we next looked for metastatic site enrichment in tissue RNA-seq data across CRPC subtypes to determine if specific CRPC subtypes are more likely to originate from a particular site. To avoid double counting, the samples were annotated as belonging to a subtype if they showed it as their major subtype (>50%). Consistent with previous studies, we find that CRPC-NE samples are more likely to be from liver (Fisher’s Exact Odds Ratio = 5.45, P = 6e-6, fig. 4) and soft tissue (Fisher’s Exact Odds Ratio = 3.55, P = 7.12e-3, fig. 4)^35,36^. While most samples from all sites are CRPC-AR (fig. S4), as expected, we do find enrichment of this subtype at lymph node (Fisher’s Exact Odds Ratio = 3.37, P = 4.97e-7, fig. 4), which is consistent with previous reports^37^.

**Fig. 4:**
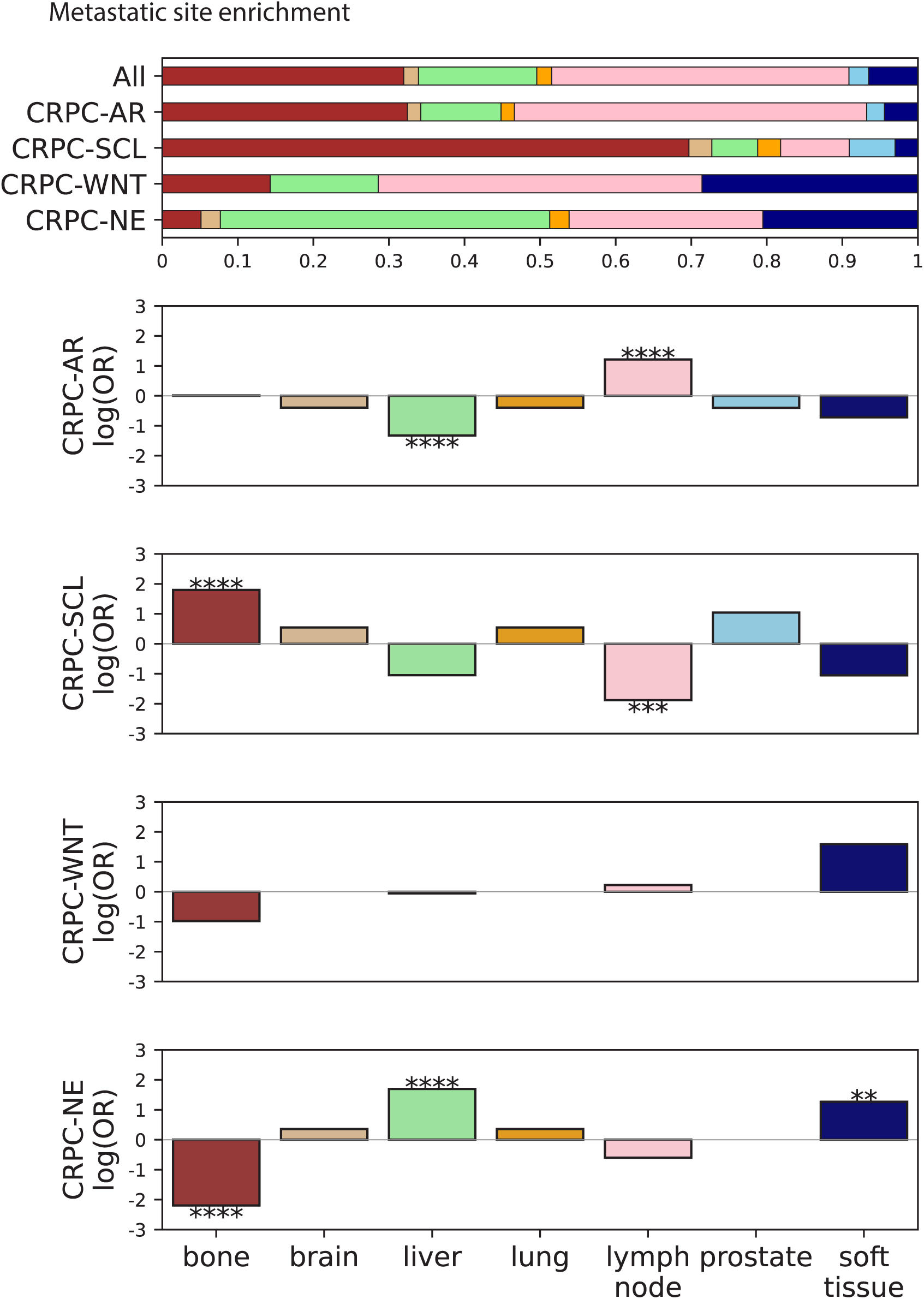
Metastatic site enrichment. Fractions of samples (across all subtypes, and for within each subtype for subtype fraction ≥0.5) from each metastatic site (top), log (natural) odds ratio (OR) of a sample coming from each metastatic site across subtype fractions (bottom).

We find that CRPC-SCL samples are more likely to originate from bone compared to other tissues (Fisher’s Exact Odds Ratio = 6.03, P = 2e-6, fig. 4). This is consistent with previous reports that PC3 (a CRPC-SCL cell line) colonizes to bone in mice^38^, and evidence of tumor cells with AP-1 signaling (that is characteristic of CRPC-SCL) in scRNA-seq from bone metastases^39^. A mechanistic study to examine how this occurs, found that protein regulator of cytokinesis 1 (PRC1), which has been implicated in promoting stemness in cancer, is elevated in prostate cancer cells with stem cell traits and drives colonization to the bone^38^. Moreover, it is well known that bone provides a unique niche for stem cells, in part because of its hypoxic microenvironment that promotes commitment to stemness^40^. Thus, we hypothesize that it is the bone microenvironment that makes it especially hospitable to CRPC-SCL, which is characterized by stem cell-like features.

### Genomic alterations of the YAP/TAZ pathway associated with epigenomic subtypes

We identified genome-wide copy number alterations (CNAs), structural variants (SVs), and single nucleotide variants (SNVs) from WGS data from cfDNA and tissue. When matched WGS from cfDNA and tissue was available, we observe a high concordance of both type and breakpoints (for SVs) between called variants, showing the reliability of calling genomic alterations from cfDNA (fig. S5). Moreover, we observe multiple events that are consistent with previous studies. Samples with CRPC-AR are enriched for amplifications of the *AR* gene (fold change = 2; binomial p value = 2.55e-5; fig. 5B, S6B); those with CRPC-NE are enriched for biallelic inactivation of *RB1*^13,17,28^ (fold change = 5.21; binomial p value = 1.41e-02, fig. S6C); and *CTNNB1* (*β*-catenin) mutations are enriched in CRPC-WNT samples (6.5% for CRPC-WNT vs. zero for others; binomial p value = 0; fig. S6D) (tables S9, S10, S11, S12, S13).

**Fig. 5:**
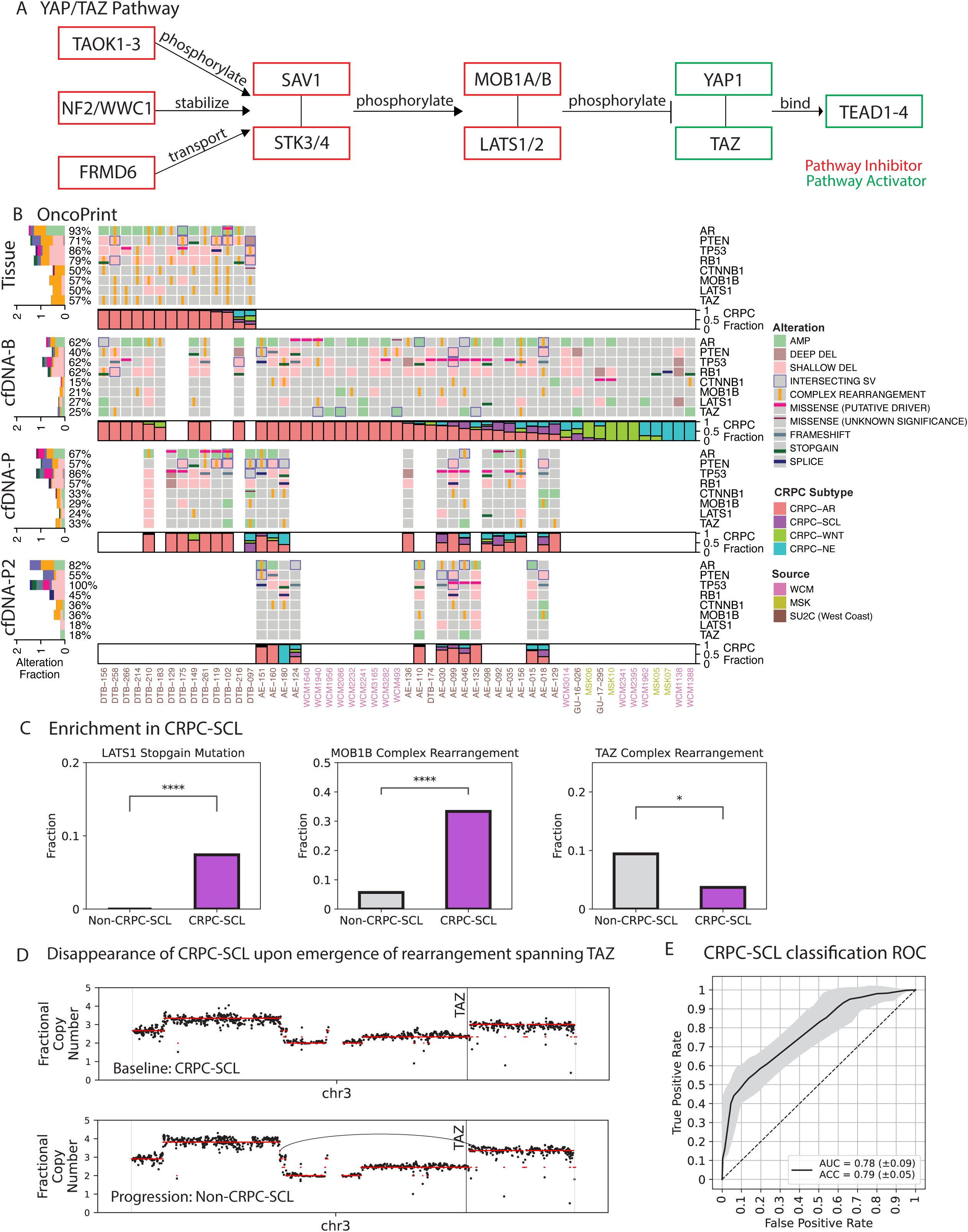
YAP/TAZ pathway alterations. (A) YAP/TAZ pathway. (B) Oncoprint shows the genomic alterations for each patient in tissue and cfDNA timepoints (where available, Baseline (B), Progression (P), and Progression2 (P2)) for various genes of interest, each row is followed by the CRPC fraction of each sample from deconvolution. (C) LATS1 stopgain mutations in CRPC-SCL, complex rearrangement of chromosomes spanning MOB1B and TAZ in CRPC-SCL. (D) AE-129 signal track along chr3 across timepoints. Fractional copy number (FCN) along chr3, TAZ gene locus marked by vertical line, location of complex rearrangement shown as curved line at Progression. (E) receiver operating characteristic (ROC) curve of logistic regression model to classify samples as CRPC-SCL.

The chromatin accessibility landscape of CRPC-SCL has been shown to be enriched for binding sites of the AP-1 and TEAD families of TFs, which work together to promote the oncogenic growth of CRPC-SCL tumors^8^. Activation of the TEAD family is dependent on the YAP/TAZ pathway (fig. 5A). At baseline, the YAP/TAZ pathway is turned off by proteins such as TAOK1/2/3, NF2, WWC1, FRMD6, SAV1, STK3/4, MOB1A/B, and LATS1/2 which work to phosphorylate, and thereby deactivate YAP1 and TAZ (an alias for WWTR1). In the absence of this deactivation mechanism, YAP1 and TAZ are in their active form, and translocate to the nucleus to bind to and activate TEAD, which in turn likely works with AP-1 to regulate the transition to CRPC-SCL^8,41^. Furthermore, YAP/TAZ target genes are upregulated in the context of CRPC-SCL^8^ (fig. S3E).

So far there have been no reports of CRPC-SCL-specific genomic alterations. YAP/TAZ pathway alterations have been reported in many other cancers, and are most prevalent in mesothelioma, kidney renal papillary cell carcinoma, and cervical squamous cell carcinoma^41^. In light of this, we sought to leverage our unique dataset to describe the genomic alterations observed in CRPC. Indeed, we find alterations of certain YAP/TAZ pathway genes enriched in samples with CRPC-SCL (fig. 5B, 5C). These include complex rearrangements of chromosome 4 spanning the YAP/TAZ pathway inhibitor *MOB1B* (fold change = 5.89, binomial p value = 1.10e-05, figs. 4B,C, S6E), and stopgain mutations in another inhibitor, *LATS1* (7.4% for CRPC-SCL vs. zero for others, binomial p value = 0, figs. 5B,C). As opposed to pathway inhibitors, we might expect activators to not be disabled in CRPC-SCL samples, and indeed we observe a depletion of a complex rearrangement of chromosome 3 spanning the YAP/TAZ pathway activator *TAZ* in these samples (fold change = 2.55, binomial p value = 4.60e-02, fig. 5B,C, S6E). In our analysis, complex rearrangements are a type of SVs defined as large events that span the entire gene or multiple genes. They are predicted to be either end-to-end telomere translocations or pericentric inversions of the region and are supported by discordant reads at the breakpoints upon manual inspection^41^ (fig. S7). We hypothesize that these events may have functional implications in regulating the genes, and thereby, the YAP/TAZ pathway.

A major advantage of using cfDNA is that it can be used for serial monitoring made feasible by its simple and minimally invasive manner of collection. Similar to the previous observation of emergence of biallelic inactivation of *RB1* accompanying the emergence of CRPC-NE in a patient (AE-180, fig. 5B)^15^, we are able to track the co-evolution of *YAP/TAZ* pathway alterations with CRPC subtype using cfDNA. The sample AE-129 began with presence of CRPC-SCL but showed reduction of this subtype at progression on ARSIs accompanied with a complex rearrangement at chromosome 3 spanning the *TAZ* gene, potentially inactivating the YAP/TAZ pathway (fig. 5B,D). Since most prostate tumors begin as AR dependent, this example where we see reduction of CRPC-SCL and increase of CRPC-AR potentially also points to the highly dynamic nature of lineage plasticity where patients may shift back and forth between subtypes and highlights the power of using cfDNA to detect such transitions.

### Hi-C data validates complex rearrangements and shows they reduce access of *MOB1B and TAZ* to their cis-regulatory elements

Large-scale chromosomal alterations have been implicated in many cancers. Loss of heterozygosity of chr8p frequently occurs in breast and prostate cancers^42,43^. Amplification of chr20q is associated with many cancer types such as pancreatic, gastric, colon, ovarian, cervical, and breast cancers^44^. Arm level deletion of chr13q, which includes the *RB1* gene, is associated with neuroendocrine prostate cancer^45^. The CRPC-SCL-enriched/depleted complex rearrangements identified in this study also represent large-scale chromosomal rearrangements. For example, the complex rearrangements spanning *MOB1B*, ranging from 9Mb to 190Mb, cover ∼48% of the chromosome on average (fig. 6B).

**Fig. 6:**
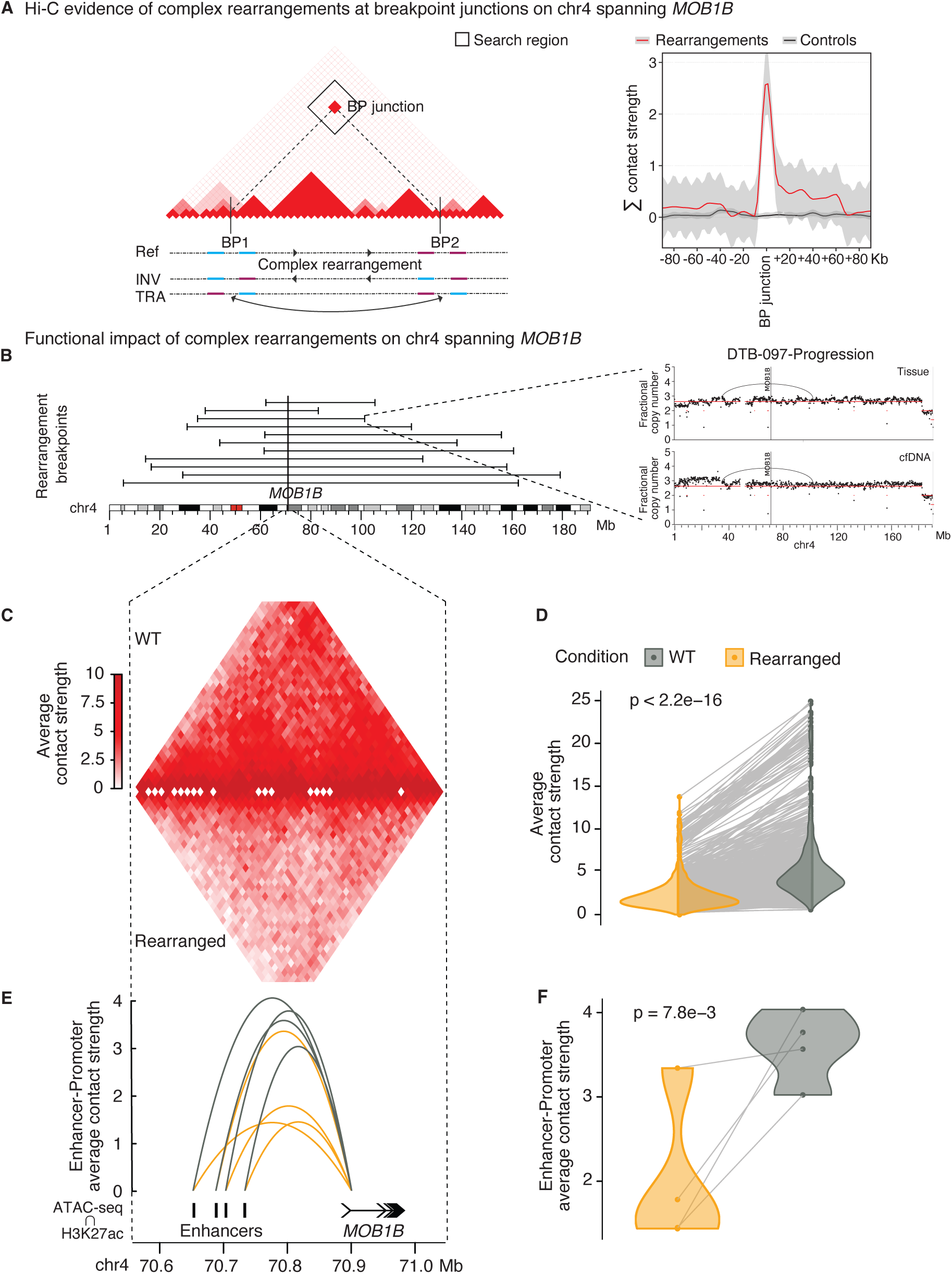
Hi-C data validates structural variants and shows complex rearrangements reduce access of *MOB1B* to its regulatory elements. (A) schematic of Hi-C contact map and indices storing the connection strength of regions surrounding example inversion (INV) and translocation (TRA) breakpoints (i.e. breakpoint (BP) junction) (left) and contact strength increase in search region relative to negative control regions support evidence of rearrangement (right). (B) breakpoint locations of complex rearrangements spanning *MOB1B* in CRPC-SCL samples (left) and example of concordant breakpoints in cfDNA and matched tissue in one sample (right). (C) contact maps in TAD surrounding *MOB1B* in CRPC-SCL samples without (wildtype (WT), top) and with (rearranged, bottom) the rearrangement. (D) Strength of the contacts averaged across samples is lower in CRPC-SCL samples with the rearrangement (rearranged) compared to those without it (WT). (E) Average interaction strength between *MOB1B* promoter and its enhancers in CRPC-SCL samples with (rearranged) and without (WT) the rearrangement. (F). Strength of the enhancer-promoter interactions averaged across samples is lower in CRPC-SCL samples with the rearrangement (rearranged) compared to those without it (WT).

Such complex SVs are characteristic of prostate cancer genomes^14,22,46^, however it is generally hard to confidently identify true positive events using short-read sequencing alone. Thus, we leveraged the matched Hi-C and tissue RNA-seq data (n=55) to evaluate 3D chromosome conformation of each sample together with its molecular state^18,47^. In the Hi-C contact map, topologically associated domains (TADs) manifest as large genomic regions with elevated internal 3D conformation interactions, whereas SVs manifest as interactions isolated to the breakpoints and their immediate surrounding regions only^18^. Thus, Hi-C data can be used to validate the SVs, besides inferring their impact on chromatin conformation^48^.

We first sought evidence for the complex rearrangements on chromosome 4 spanning *MOB1B* and enriched in CRPC-SCL samples from matched Hi-C data. Towards this end, we looked at the contact strength in the Hi-C map at the locus that stores the interaction between the two breakpoints of the complex rearrangement, referred to as the breakpoint junction (fig. 6A). Assuming the event is true, we expect increased interactions at the breakpoint junction relative to surrounding regions, since regions that are far away from each other in the reference genome are now close by in the tumor genome due to the rearrangement. Indeed, we observe higher contact strength at the breakpoint junction and a reduction of contact strength in its surrounding bins, whereas randomly selected control regions of the same length show no such increase (fig. 6A). Similarly, the complex rearrangements enriched in non-CRPC-SCL samples on chromosome 3 spanning *TAZ* are also supported by increased interactions at breakpoint junctions in those samples (fig. S8A). Furthermore, concordant breakpoints identified separately in matched tissue and cfDNA provide further support for these rearrangements (fig. 6B,C).

We next investigated a potential functional role of the rearrangements in regulating CRPC-SCL. We expected that genes affected by complex rearrangements may have decreased access to their cis-regulatory elements such as enhancers since *MOB1B* shows lower expression in CRPC-SCL samples relative to non-CRPC-SCL (fig. S3E). We analyzed 0.5 Mb region spanning *MOB1B* and find that the strength of 3D contacts, averaged across samples, is lower in CRPC-SCL samples with the rearrangements when compared to those without it (fig. 6B,C,D). By overlaying H3K27ac and ATAC-seq marks with Hi-C contacts, we find that CRPC-SCL samples with the rearrangement show reduced number and contact strength for enhancers connected to *MOB1B* promoter (fig. 6E,F). These trends were reproduced for *TAZ* in non-CRPC-SCL samples (fig. S8). These findings suggest that the complex rearrangements may be associated with *MOB1B* and *TAZ* downregulation at the transcriptional level by reducing promoter access to their enhancers, in addition to their regulation by phosphorylation at the proteomic levels^41^. Even though the breakpoints of rearrangements are far from *MOB1B* and *TAZ* promoters and enhancers, previous studies have shown that large SVs may have a long-distance effect through their impact on the 3D spatial architecture of the chromatin^49^. In line with this, we expect that these rearrangements may also be associated with disruption of other TADs on the chromosome besides the ones surrounding the genes in question. Indeed, by using samples without presence of CRPC-SCL or complex rearrangements as reference, we find that CRPC-SCL samples with the rearrangements have overall fewer conserved TADs than the samples without the rearrangements (fig. S8B).

### Logistic regression model to predict CRPC-SCL using genomic alterations

Besides the *MOB1B*, *TAZ*, and *LATS1* variants discussed above, we checked if overall the genomic alterations in the YAP/TAZ pathway are subtype-specific enough to enable identification of CRPC-SCL subtype. Towards this end, we trained a logistic regression model to classify samples containing CPRC-SCL based on alteration events in the YAP/TAZ pathway. The input to the method is the presence or absence of genomic alterations for each gene of the pathway and the output is the probability that the sample exhibits presence of CRPC-SCL subtype. Using Monte Carlo Cross Validation and L1 regularization with a manual parameter search to find the optimal number of features and lambda value for each fold, we classified samples as CRPC-SCL with an accuracy of 0.79 and area under the receiver operating characteristic curve (AUROC) of 0.78, averaged across folds (fig. 5E). These results show that these genomic events are predictive of CRPC-SCL.

### CRPC subtypes and genomic heterogeneity and instability

Chromosomal instability and heterogeneity, measured by loss or rearrangement of chromosomes has been reported as a hallmark of cancer and is associated with high disease burden, treatment failure, metastatic progression, and death^50^. Chromosomal instability can be described numerically (i.e. the number of copies of whole chromosomes) or structurally (i.e. the fraction of each chromosome altered)^51,52^. Here, we quantify chromosomal instability as the fraction of the genome altered (FGAlt) by a copy number or structural variant event. We observe that CRPC-SCL samples are enriched for very high FGAlt (fig. 7A,7B). It is possible that high FGAlt may partly contribute to the shorter survival of patients with CRPC-SCL, as it is also associated with aggressive disease in castration-sensitive prostate cancer^53^ (fig. 2C).

**Fig. 7:**
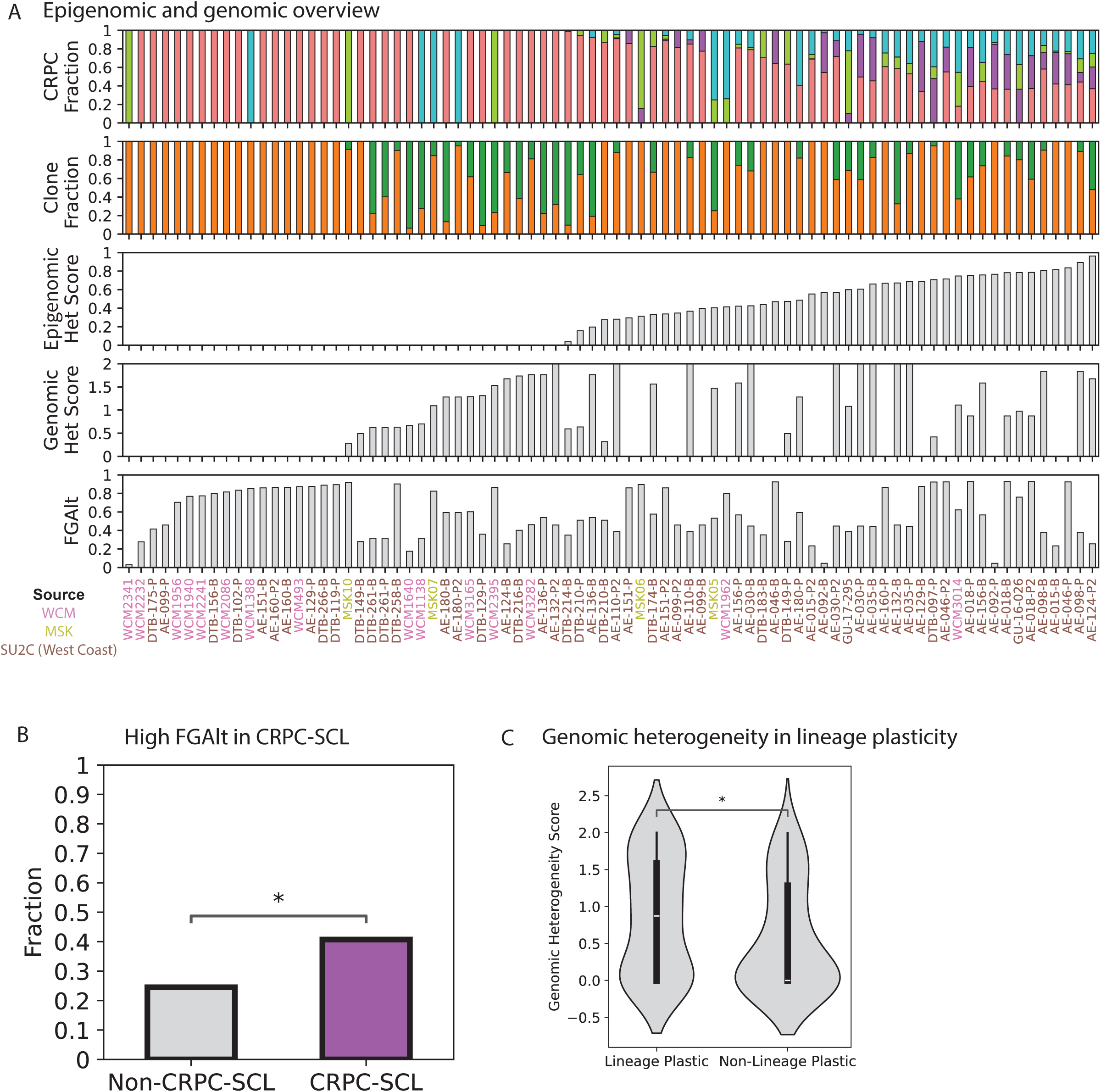
Genomic heterogeneity and instability and CRPC subtypes. (A) Per-sample molecular and genomic overview: CRPC fraction from deconvolution (top row), tumor clone fraction composition (second row), molecular heterogeneity score (third row), genomic heterogeneity score (fourth row), and fraction of genome altered (last row). (B) High FGAlt enrichment in CRPC-SCL samples. (C) Genomic heterogeneity score distribution in lineage plastic and non-lineage plastic samples.

We have shown that samples with lineage plastic subtypes tend to be more heterogeneous for the presence of multiple subtypes (fig. 3). Because many genomic events are early cancer events and are associated with different molecular states, we expect genomic and epigenomic heterogeneity to relate. Epigenomic heterogeneity refers to the diversity of CRPC subtype composition within a given sample. Genomic heterogeneity refers to the diversity in tumor clone composition within a given sample. Genomic clones are estimated for each sample by HATCHet, the clone-specific copy number caller we used to estimate genome-wide CNAs^54^. CNAs are especially useful in identifying genomic clones because they occur at large scales, often affecting considerable genomic regions, and can be called in an allele-specific manner. We used clone-specific CNAs to construct a genomic heterogeneity score to account for the number of tumor clones, as well as the similarity between them. The score is defined as *n* × (1 − *JI*), where *n* is the number of tumor clones, and *JI* is the Jaccard index between clones. We find that lineage plastic samples also show higher genomic heterogeneity scores, which is positively correlated with the number of epigenomic subtypes (Spearman R = 0.22, P = 2.37e-02, fig. 7A,C). Because just the number of subtypes may not fully capture the epigenomic heterogeneity, we assigned an intra-sample epigenomic heterogeneity score to each sample that measures the diversity of the CRPC fraction composition. The score is defined by the normalized Shannon entropy of the subtype fractions, such that a sample entirely comprised of one CRPC subtype would have a score of 0, and a sample comprised of equal parts of all four CRPC subtypes would have a score of 1. Indeed, we observe that genomic heterogeneity is positively correlated with epigenomic heterogeneity (Spearman R = 0.21, P = 3.36e-02).

## Discussion

In this study we annotate the epigenomic subtypes of CRPC patient samples using nucleosomal profiling (cfDNA) and transcriptomic (tissue) data. We find that the histologic classifications of adenocarcinoma and neuroendocrine do not capture the intra- and inter-sample subtype heterogeneity, potentially missing important molecular information relevant for therapeutic decisions. Overall, we find that higher subtype heterogeneity is also associated with higher genomic heterogeneity, and CRPC-SCL samples tend to show higher genomic instability as measured by FGAlt. Enrichment of CRPC-SCL tumors in bone biopsies highlights the importance of studying the tumor immune microenvironment of this subtype in future since bone is the most common site of prostate cancer metastasis. We validate our subtype fraction estimations with scRNA-seq and confirmed that non-tumor cells do not confound our calculations^8,30,39^. By relating the genomic events from WGS with the epigenomic subtypes, we find novel genomic alterations at the YAP/TAZ pathway genes that are associated with the CRPC-SCL subtype. The breakpoints of these rearrangements are validated by Hi-C reads. Indeed, by combining all putative functionally disrupting events in the YAP/TAZ pathway, we are able to predict the presence of CRPC-SCL in a given sample with a high accuracy of 0.79 using a logistic regression model. Previously, few genomic alterations such as *RB1* loss were known to be associated with lineage plasticity in CRPC, and our study adds more genomic events to this list. Interestingly, we find the regression model for CRPC-SCL prediction trained on cfDNA WGS samples outperforms the one trained with tissue samples (RNA-seq and WGS), potentially due to higher fraction of cfDNA samples that exhibit high fraction of CRPC-SCL in cfDNA in our cohort.

It is well known that alterations affecting non-coding regions of the genome (e.g. enhancers) can play a major role in cancer^55^. In prostate cancer, *AR* enhancer amplifications and fusion of the 5’ *TMPRSS2* UTR to *ERG* are well studied^14,56^. Large complex rearrangements that span the entire gene or multiple genes have generally been harder to interpret. The availability of matched Hi-C data from patient samples with RNA-seq and WGS allowed us to analyze the enhancer— promoter contacts in samples with versus without rearrangements. Our findings point to potential dysregulation of *MOB1B* by rearrangements on chromosome 4 associated with presence of CRPC-SCL, and of *TAZ* on chromosome 3 associated with absence of CRPC-SCL. While Hi-C data provides support for putative functional roles of these SVs, future studies will be needed to establish the mechanism of their action of gene disruption as well as their role in up- or down-regulation of the YAP/TAZ pathway and lineage plasticity.

We have shown that both tissue RNA-seq and cfDNA WGS can be used to infer the molecular CRPC subtypes and generally provide consistent results. This is especially impactful since cfDNA is minimally invasive and can be used for serial monitoring. Interestingly, unlike previous observations for genomic heterogeneity where cfDNA exhibits more heterogeneity than tissue, we generally did not observe this trend for epigenomic subtype heterogeneity (fig. S9A)^15^. This observation is also supported by another work using spatial gene expression profiling in which the authors found concordance in gene expression and phenotype classification between metastases from the same individual, albeit with exceptions^11^. However, this observation of absence of higher epigenomic heterogeneity in cfDNA relative to tissue could also be partly due to the limitations of using cfDNA for mixed subtypes inference. Thus, we estimated the limit of detection of the current computational approach to infer CRPC subtype fractions from cfDNA. Under the assumption that each metastatic site sheds cfDNA from molecular subtypes in proportion to their presence in tissue, we find that a subtype fraction of ≤ 0.2 in tissue cannot be detected in cfDNA (fig. S9). These metrics can serve as a guide for the clinical use of cfDNA WGS for molecular subtype inference and guide the further development of computational approaches for subtype inference. Overall, our study provides a framework to move beyond histological classifications of patient tumors to epigenetic subtyping using a clinically feasible assay of cfDNA WGS and shows that it can be used beyond biomarker for discovery of novel genomic alterations associated with lineage plasticity.

## Methods

### Cohort Overview

We sequenced cfDNA from 20 patients with CRPC. By combining this with published data, we have amassed a cohort of 488 patients with metastatic CRPC. cfDNA and tissue were collected from 68 and 455 patients, respectively, 23 of which were time matched (fig. 1A) (table S1)^8,14,15,18,19,47^.

We analyzed cfDNA WGS from 95 cfDNA samples from 68 patients. 61 samples were from a previously published study^15^, which were enriched for adenocarcinoma patients with only 2 patients showing neuroendocrine histology. Thus, we sequenced 20 patient samples from WCM and MSK, out of which 8 showed neuroendocrine histology. For some of these patients, we also generated RNA-seq (n=8) and WGS (n=5) data from matched tissue. Additionally, we sequenced 14 samples from patients with no cancer, as controls. cfDNA samples in Herberts et al. were collected from patients at clinically relevant timepoints, such that patients had either 1 (n = 14), 2 (n = 13), or 3 (n = 7) cfDNA samples in total^15^. Tumor fraction ranged from 0.11 to 0.92 across samples (mean = 0.46, median = 0.45)^57^. Mean coverage achieved across samples ranged from 8.64X to 182.39X (mean = 87.57X, median = 102.17X) (fig. 1B) (table S1). 32,444 structural variants (SVs) were identified across all samples, with a mean of 421.35 SVs per sample (range: 17 – 1,086). 2,165,361 single nucleotide variants (SNVs) were identified across all samples, with a mean of 5,217 non-intergenic, annotated SNVs per sample (range: 15 – 59,738).

Tissue RNA-seq, WGS, and Hi-C datasets from 482, 105, and 55 samples, respectively were analyzed ^8,14,18,19,21^. 514, 71, and 327 samples from patients with adenocarcinoma, neuroendocrine, and poorly differentiated/inadequate for diagnosis/not available histologies, respectively. Out of these 123 samples were from WCM and 789 were from other published studies^8,14,18,19,47^. Tissue samples were collected from various metastatic sites including soft tissue (n = 71), lymph node (n = 336), bone (n = 263), bladder (n = 2), prostate (n = 19), lung (n = 15), brain (n = 9), liver (n = 122), skull base (n = 1), ureter (n = 1), adrenal (n = 3), mediastinum (n = 1), spine (n = 1), epidural thoracic (n = 1), and intercostal (n = 1) (table S1).

### cfDNA WGS

#### Patient samples from WCM and MSK

Fresh blood or frozen plasma samples from consented prostate cancer patients were collected under Weill Cornell Medicine Institutional Review Board approval 20-04021962 prior to sample acquisition. Patient blood was collected in Streck tubes and spun at 1600xg for 15 min at 25°C. The plasma was decanted from the top fraction of the Streck tube and cfDNA was isolated immediately if possible, or frozen at −80°C. Plasma cfDNA was isolated using the NeoGeneStar cfDNA variable volume kit according to manufacturer’s instructions. For germline DNA, we took the top 2ml of the remaining red blood cell component of the sample from the Streck tube, added 4ml RPMI-1640, mixed gently, underlayered with 3ml Histopaque 1.077, spun 400xg for 30 min., room temperature, with no break, and isolated the peripheral blood mononuclear cells (pbmcs) from the interface. DNA from the pbmcs was isolated using the Qiagen DNeasy Blood and Tissue Kit according to manufacturer’s instructions. All samples were quantitated on Qubit HS and frozen at −80°C. Library preparation and WGS were performed at the New York Genome Center using the Kapa Hyper PCR+ cfDNA on the Illumina NovaSeq with 150 paired-end reads and coverages of 0.1, 10, 40 or 80X. Library quality was verified using Agilent Bioanalyzer 2100 and Qubit.

The raw data was aligned to hg38 using BWA mem (version 0.7.15) with parameters -Y -K 100000000 -t 8. Base quality scores were adjusted using GATK ApplyBQSR (version 4.1.0.0).

#### Publicly available patient samples

Raw data was downloaded from the European Genome-Phenome Archive (EGAS00001005783). Fastqs were aligned to hg38 using BWA mem (version 0.7.17) with parameters -M -t 20.

#### Publicly available PDX samples

Raw data was downloaded from NCBI BioProject (PRJNA900550)^10^. Fastqs were aligned to hg38 and mouse reads were removed using the same pipeline described in De Sarkar and Patton et al^10^.

### ATAC-seq

#### Human PBMCs

Fresh PBMCs were collected from a healthy control WCM sample and processed according to the Omni-ATAC protocol in Corces et. al^58^. The partially tagmented samples were sent to the WCM Epigenomics Core for the remainder of the library prep (Illumina) and P2 100 cycles run on NextSeq2000.

Fastqs were aligned to the hg38 reference using BWA mem (version 0.7.15) with parameters -t 6 -M -a -h 100 -L 50,50 and -O 7. Open chromatin peaks were called using genrich (version v0.6.1, https://github.com/jsh58/Genrich) with parameters -j -d 50 -r -e chrM -p 0.01 -q 0.05 -a 20 -g 20 -l 100 -v. IDR (version v2.0.4.2, https://github.com/nboley/idr) was used to get consensus peaks across technical replicates with parameters --output-file-type narrowPeak -- rank p.value --plot --use-best-multisummit-IDR --verbose.

#### Publicly available mouse PDX samples

Publicly available LuCaP ATAC-seq data was downloaded from GEO (GSE156292)^10^. Fastqs were aligned to hg38 using bowtie2 (version 2.4.1) with parameters --wrapper basic-0 -k 10 -t - p 16. Open chromatin peaks were called using genrich (version 0.6.1) with parameters -t -j -v -r -q 0.05 -l 100 and -E to exclude gaps, patches, centromeres, alternative haplotypes, centromeres, and black-listed regions.

### RNA-Seq

Publicly available tissue RNA-seq was downloaded directly from Tang et al.^8^, https://quigleylab.ucsf.edu/data^14,47^, and https://github.com/cBioPortal/datahub/raw/refs/heads/master/public/prad_su2c_2019^19^.

### Tumor fraction

ichorCNA (version 0.3.2, https://github.com/broadinstitute/ichorCNA) with default parameters was used to estimate tumor fraction from WGS cfDNA^57^.

### Copy number alterations

#### cfDNA WGS and matched tissue WGS

HATCHet was used to call copy number alterations from WGS cfDNA and matched tissue (when available)^54^. This applied to the in-house (WCM and MSK) and Herberts, Annala, and Sipola et al^15^ cohorts.

#### Tissue WGS

Copy number alterations were called using CopyCat and downloaded directly from https://quigleylab.s3.us-west-2.amazonaws.com/datasets/2018_04_15_matrix_CN_integer_symbol_copycat.txt.zip. This applied to the SU2C (West Coast) cohort^14^.

### Structural variant calling

#### cfDNA WGS

SVs from publicly available data were retrieved from the authors^15^. To achieve symmetry, we followed the protocol of Herberts et al for our in-house samples (relaxing certain thresholds to match sequencing depth), using Breakfast (version 0.6.0, https://github.com/annalam/breakfast) with parameters --max-frag-len = 1000 and --anchor-len = 30, requiring at least 5 unique junction-spanning reads^15^. The following thresholds were used to further filter the data: at least 4 mutant reads, at least 4 reads of total depth, VAF ≥ 0.1, average mapping quality of at least 20, and average sidedness (distance to read end) of at least 15.

#### Tissue WGS

SVs from publicly available data were called using Lumpy and were downloaded directly from https://s3-us-west-2.amazonaws.com/feng.genomics/WGBS/secondary_data.zip^18^. SVs found in more than 4 samples (breakpoint within 10 bps) in the cohort were considered non-unique and removed. SVs that were found in either of two databases of germline variants (dbVar^59^ and InvFEST^60^) were removed (breakpoints within 100 bps).

### Single nucleotide variant calling

#### cfDNA WGS

SNVs from publicly available data were retrieved from the authors^15^. To achieve symmetry, we followed the protocol of Herberts, Annala, Sipola et al for our in-house samples, using mutato (version 0.9.0, https://github.com/annalam/mutato) with arguments --alt-reads = 5 --alt-frac = 0.05. Only SNVs with at least 1 supporting read with average mapping quality ≥ 10 and within 15 bp of the read end, VAF ≥ 0.1, and a population allele frequency < 0.5% (from GNOMAD, version 3.0) were retained^15^. The functional impact of the SNVs was annotated using ANNOVAR (version 20191024) with parameters hg38 regGene gene-based annotation^61^.

#### Tissue WGS

SNVs from publicly available data were called using Strelka and were downloaded directly from https://quigleylab.s3.us-west-2.amazonaws.com/datasets/2018_04_15_WCDT_somatic_vcf.tar^14,18^. The functional impact of the SNVs was annotated using ANNOVAR (version 20191024) with parameters hg38 regGene gene-based annotation^61^.

### CRPC subtype-specific ATAC-seq peaks

ATAC-seq peaks in CRPC cell lines and organoids were called using genrich (version 0.6.1) with parameters -t -d 50 -e chrM -j -v -r -a 20 -g 20 -q 0.05 -l 100 and -E to exclude gaps, patches, centromeres, alternative haplotypes, centromeres, and black-listed regions. IDR (version 2.0.2) was used to get consensus peaks across replicates with parameters --output-file-type narrowPeak --rank p.value --verbose. Differentially accessible ATAC-seq peaks (DAAPs) generated with macs2 peaks were downloaded from Tang, Xu, Wang, & Wong et al^8^. For every macs2 peak of CRPC subtype A, if there was at least one cell line or organoid of subtype A with an overlapping genrich peak and there were no cell lines or organoids of any other subtype with an overlapping genrich peak, the subtype specific peak was defined by a start (min(start of all genrich peaks of cell lines/organoids of subtype A that overlap the macs2 DAAP)), end (max(end of all genrich peaks of cell lines/organoids of subtype A that overlap the macs2 DAAP)), and summit (int(median(summits of all genrich peaks of cell lines/organoids of subtype A that overlap the macs2 DAAP))). Any peak that overlapped a genrich ATAC-seq peak of our in-house no cancer control sample (ATAC-seq Human PBMCs above) was removed. Overall, there were 11,658 CRPC-AR sites, 13,959 CRPC-SCL sites, 13,732 CRPC-WNT sites, and 11,642 CRPC-NE sites (table S14).

### CRPC subtype fraction estimation

#### cfDNA WGS

We performed deconvolution using Keraon^10^. The signature matrix was built by averaging the WGS cfDNA mean coverage across PDXs of the same CRPC subtype in CRPC subtype-specific accessible chromatin sites of that subtype (table S5). Because we had no CRPC-WNT PDXs, we used a CRPC-WNT patient for the signature and excluded it from other analyses. Features used were central coverage (30 bp window centered at the summit) and window coverage (990 bp window centered at the summit). Signatures were also computed from 14 healthy control samples and were subtracted out of the PDX signature (tables S5,S6). The mixture matrix was the WGS cfDNA mean coverage of each sample in CRPC subtype-specific accessible chromatin sites (fig. S1B).

#### Tissue RNA-seq

Deconvolution was performed using EPIC^20^. The signature matrix was built based on averaging the expressions (Transcripts Per Million, TPM) across cell lines and organoids of the same CRPC subtype in CRPC signature genes^8^ (tables S2,S3). The mixture matrix was the gene expression values (TPM) of each sample for CRPC signature genes^8^. The mixture matrix is then regressed back onto the signature matrix to estimate fractional contributions of each CRPC subtype to each patient’s expression profile (fig. S1A).

### scRNA-seq analysis

Single-cell RNA-seq count matrices derived from human metastatic CRPC biopsy samples were downloaded directly from GEO (GSE210358)^9^. To perform pseudo-bulking and conversion to TPM, gene counts were summed across all cells in each sample (only common genes across samples were retained). Gene lengths were acquired from a biomaRt query to use in TPM conversion^62^. For tumor-only pseudo-bulking, the same approach was used across only tumor cells (GSM6428952)^9^. To annotate these cells, the authors used a hierarchical strategy wherein broad categories of epithelial, immune, and stromal cells were identified using expression of marker genes for these categories. Expression of genes that are known to be upregulated in prostate cancer are used to identify tumor cells. Cell type assignments were confirmed by showing inferred CNVs were higher in tumor cells relative to non-tumor cells^30^.

### Metastatic site enrichment

For each CRPC subtype, we performed a two-tailed Fisher’s exact test to determine if the odds of samples from that subtype coming from a particular metastatic site versus others were statistically significant. This analysis was performed on the tissue dataset and was limited to samples with CRPC subtype fraction ≥ 0.5.

### Genomic event enrichment

For each comparison, we performed a one-tailed binomial test to determine if the probability of samples from a given group having the alteration was significantly different from the probability of other samples having the alteration. We had segment-level copy number calls for cfDNA and gene-level scalar copy number calls for tissue. For AR amplifications, we checked for amplifications (CN>3 in cfDNA, CN>3 in tissue – values chosen based on CN distribution) in CRPC-AR samples versus others. For biallelic inactivation of RB1, we checked for homozygous deletions (CN≤1.2 in cfDNA, CN≤0.5 in tissue – values chosen based on CN distribution) or a combination of at least two of the following events: heterozygous deletions (1.2≤CN<1.75 in cfDNA, 0.5≤CN<1.6 in tissue – values chosen based on CN distribution), inactivation by SV (i.e. SV breakpoint intersecting gene), stopgain, and splice mutations. For CTNNB1 mutations, they were putative driver missense mutations. Only complex rearrangements that were significant in both cfDNA and tissue were reported.

### CRPC-SCL prediction using genomic variants

We trained a logistic regression model to classify samples as CRPC-SCL or not, using a 2:1 train/test split. We performed this 10 times using different seeds each time. We performed L1 regularization with a manual parameter search to find the optimal number of features and lambda value, testing c values within an np.logspace(−4,4,20) for each train/test split. The feature matrix had samples in the rows and YAP/TAZ pathway genes in the columns and was populated by 1 if the sample had events deemed putatively functional (a complex rearrangement, missense (putative driver) mutation, or stopgain mutation of the gene), and 0 otherwise. The target classes were 0 if the sample was not CRPC-SCL and 1 if it was. We checked that including other genomic events did not significantly change the performance of the model.

We also trained models to classify the other CRPC subtypes, but CRPC-SCL performed the best by far. We hypothesize that classification of CRPC-NE (AUC=0.61, ACC=0.65) and CRPC-WNT (AUC=0.64, ACC=0.64) is better than random because many of the samples also contain CRPC-SCL.

### Hi-C analysis

Normalized contact maps were downloaded from GEO (GSE249494)^18^. We leverage the information obtained from the Hi-C experiment in two main tasks. First, the predicted breakpoints were validated using Hi-C interactions by searching the nonlinear contact map. We compare the contact strength at the breakpoint junction to its surrounding regions. We define a search space of 10 Hi-C bins (100 Kb) on each side of the breakpoint junctions. For each sample with the complex rearrangement and the Hi-C contact map available, we calculated the contact values in the search space, aggregated across samples. Then, we evaluate the trend of the aggregated contact strength in the search space by fitting a curve using a smoothing window of two Hi-C bins (20 Kb). As a control, we evaluate the same trends in random regions to show that nonlinear interactions at the same distance are only seen due to breakpoints (fig. 6A). Secondly, we assessed the functional effect of the rearrangements on the Hi-C interactions at genomic regions associated with the CRPC-SCL subtype transition and the impact on enhancer-promoter interactions. We evaluated the average strength per contact within 500 Kb TADs harboring the YAP/TAZ inhibitor, MOB1B, using CRPC-SCL samples with and without the rearrangement (fig. 6C,D). We reproduced these trends in YAP/TAZ activator, TAZ, in Non-CRPC-SCL samples (fig. S8). Enhancer regulatory regions were then defined by the overlap of in-house generated ATAC-seq peaks^8^ and DU145 H3K27ac marks downloaded from https://figshare.com/articles/dataset/H3K27ac_Signal_Track_for_DU145/12442796. Thus, we further verified the specific change in the average strength of enhancer-promoter connections between SCL samples with and without the rearrangements (fig. 6E,F, S6C).

### Epigenomic and genomic heterogeneity scores

The epigenomic heterogeneity score is the Shannon entropy of the subtype fractions, normalized by the number of possible CRPC subtypes (i.e. 4) to restrict the range from 0 to 1 (table S15).

The genomic heterogeneity score is # *genomic clones* × (1 − *Jaccard index between clones*) where the Jaccard index was 1 for samples with only one clone (table S15).

### Fraction of genome altered

Fraction of genome altered is the sum of CNA segment lengths with abnormal copy number in at least one clone intersected with SVs (excluding interchromosomal translocations) divided by the length of the genome (table S16). We find that high FGAlt (>0.85) is enriched in CRPC-SCL. This trend is the same across thresholds (fig. S10). We find that CNAs are the largest contributor to FGAlt.

### Survival analysis

We generated Kaplan-Meier curves using progression free survival in units of days as the measure of time. Every event was counted as progression. We performed a one-tailed log rank test to determine if the curves were statistically different from one another (fig. 2C).

### Limit of detection

We compared CRPC subtype fractions in tissue RNA-seq and cfDNA WGS in patients for whom we have matched data (fig. S9A). We performed in-silico simulations to estimate our limit of detection with respect to the fraction of CRPC subtype present, the tumor fraction of the sample, and the average sequencing coverage of the sample.

#### Subtype fraction

To estimate the lowest fraction of CRPC subtype detectable from cfDNA, we chose patient AE-180 as a model. At baseline, we estimated he was 100% CRPC-AR (tumor fraction = 0.4718, mean coverage = 111.1X). At progression2, however, he was 100% CRPC-NE (tumor fraction = 0.614, mean coverage = 69.67X). By mixing specific amounts of AE-180-Baseline with AE-180-Progression2 and a healthy control, we were able to create a sample with a target CRPC-AR fraction, while maintaining a fixed coverage (40X) and tumor fraction (0.4718), the tumor fraction of AE-180-Baseline without mixing. For each target CRPC-AR fraction, we repeated the experiment 100 times, each with a different seed, generating a distribution of estimated CRPC-AR fractions across experiments. By assuming the target CRPC-AR fraction as ground truth, we evaluated our statistical and systematic errors in estimating subtype fractions. The statistical error was never more than 2.1% across target CRPC-AR fractions (fig. S9B). When the CRPC subtype fraction is less than 0.3, we do not detect it at all. When it is close to this limit, our systematic error is high, and we underestimate the contribution of the small subtype.

We compute the fractions of AE-180-Baseline, AE-180-Progression2, and control required for mixing to achieve the desired target CRPC-AR and CRPC-NE fraction as follows: We define three samples *S*_1_, *S*_2_, and *S*_3_ with CRPC-AR fraction *F*_1_, *F*_2_, and *F*_3_, tumor fraction *T*_1_, *T*_2_, and *T*_3_ and coverage *C*_1_, *C*_2_, and *C*_3_ such that *T*_1_ ≠ *T*_2_ ≠ *T*_3_ and *C*_1_ ≠ *C*_2_ ≠ *C*_3_. Let *S*_3_ be the healthy sample such that *T* = 0. Our target mixed sample is *S*_4_, such that 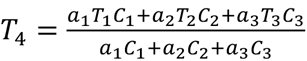 and *C*_4_ = *a*_1_*C*_1_ + *a*_2_*C*_2_ + *a*_3_*C*_3_, for some constants *a*_1_, *a*_2_, and *a*_3_. This relies on the assumption that every sample is made up of some (potentially zero) tumor and healthy component such that 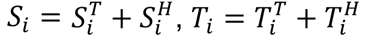, and 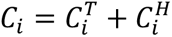. Then 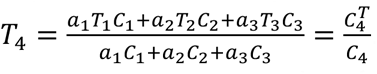. And *a*_1_*C*_1_ + *a*_2_*C*_2_ + *a*_3_*C*_3_ = *C*_4_, *a*_1_*T*_4_*C*_1_ + *a*_2_*T*_4_*C*_2_ + *a*_3_*T*_4_*C*_3_ = *a*_1_*T*_1_*C*_1_ + *a*_2_*T*_2_*C*_2_ + *a*_3_*T*_3_*C*_3_, and *C*_1_(*T*_4_ − *T*_1_)*a*_1_ + *C*_2_(*T*_4_ − *T*_2_)*a*_2_ + *C*_3_(*T*_4_ − *T*_3_)*a*_3_ = 0. Also, (*a*_1_*T*_1_*C*_1_ + *a*_2_*T*_2_*C*_2_ + *a*_3_*T*_3_*C*_3_)*F*_4_ − (*a*_1_*T*_1_*C*_1_*F*_1_ + *a*_2_*T*_2_*C*_2_*F*_2_ + *a*_3_*T*_3_*C*_3_*F*_3_) = 0, and (*a*_1_*T*_1_*C*_1_)(*F*_4_ − *F*_1_) + (*a*_2_*T*_2_*C*_2_)(*F*_4_ − *F*_2_) + (*a*_3_*T*_3_*C*_3_)(*F*_4_ − *F*_3_) = 0. Thus, we solve the following linear equation to solve for fractions *a*_1_, *a*_2_, and *a*_3_:

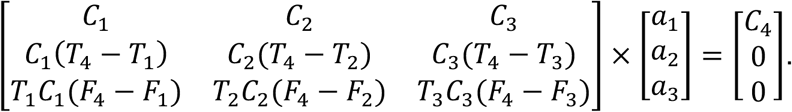

Statistical error was estimated by the standard deviation of CRPC-AR fraction across 100 runs of a given target (CRPC-AR, CRPC-NE) fraction pair. Systematic error was estimated by the difference between the mean CRPC-AR fraction across 100 runs of a given target (CRPC-AR, CRPC-NE) pair and the actual CRPC-AR fraction of the sample without mixing.

#### Tumor fraction and coverage

To estimate our ability to accurately detect CRPC subtype fraction from cfDNA with respect to coverage and tumor fraction, we chose sample AE-092-Baseline (coverage = 17.18X, tumor fraction = 0.78), who had a CRPC-SCL fraction of 0.43, as a model. We chose this sample because of its high tumor fraction and relatively high CRPC-SCL fraction. By downsampling and mixing specific amounts of AE-092-Baseline with a healthy control, we were able to create a sample with a target coverage and tumor fraction combination. For each target coverage, tumor fraction pair, we repeated the experiment 100 times, each with a different seed, generating a distribution of estimated CRPC-SCL fractions across experiments. By assuming the CRPC-SCL fraction without downsampling or mixing – i.e. 0.43 – as ground truth, we evaluated our statistical and systematic errors in estimating subtype fractions. Statistical error stems from having fixed, finite coverage and can be estimated by the standard deviation of CRPC-SCL fraction estimates across the 100 experiments. Although the statistical error goes up with lower coverage and tumor fraction, it remains below 5% for coverage down to 3X and tumor fraction down to 0.1. Using these experiments as a template, we estimated the statistical errors on each of our patient samples using their coverage and tumor fraction. On average, the statistical error of the cohort was 1.3% (fig. S9C). We find that the systematic error is mostly independent of coverage but increases with lower tumor fraction and remains around 6% or below for tumor fraction down to 0.1. Using these experiments as a template, we estimated the systematic errors on each of our patient samples using their coverage and tumor fraction. On average, the systematic error was 1.5% (fig. S9D). Calculations were performed similarly to estimating limit of detection of subtype fraction above.

## Data Availability

Processed data is provided in supplemental tables. Raw data will be made available after publication.

## Acknowledgements

We thank the Khurana and Chen lab members for their input and discussion. We are grateful to the Englander Institute for Precision Medicine for WCM CRPC patient samples, including Alexandria Norton, Christina Federov, John Mills, and John Otilano for their facilitation. We also thank the WCM Genomics Core and Epigenomics Core facilities.

## Funding

S.EN. thanks the National Institutes of Health (R25CA233208) for their support. M.R. thanks the National Institutes of Health (T32 GM083937) for their support. T.A.G. was supported by a Medical Scientist Training Program grant from the National Institute of General Medical Sciences of the National Institutes of Health under award number: T32GM152349 to the Weill Cornell/Rockefeller/Sloan Kettering Tri-Institutional MD-PhD Program. E.K. thanks the National Institutes of Health (R01CA218668 and P50CA211024), the Department of Defense (HT9425-23-1-0074), the Ann and William Bresnan Foundation, and the WorldQuant Foundation for their support.

## Author Contributions

M.R., A.M.F., and E.K. conceived of and designed the project. M.R., A.M.F., and E.K. wrote the manuscript with the help of all authors. M.R. and A.M.F. performed the majority of the data analysis. S.C. performed cfDNA sample preparation for WGS. M.M., M.B. and B.J.R. redesigned HATCHet for cfDNA WGS. W.L., A.B.T., T.A.G., S.EN., R.B., P.D., and K.G. helped with data analysis. J.M.M., B.D.R., M.A., and M.AA. performed pathological review. M.S., J.M., S.T., D.N., A.M., J.N., C.N.S., J.M., and A.S. managed and coordinated WCM data and sample delivery. E.B., H.I.S., and Y.C., managed and coordinated MSK data and sample delivery. B.J.R. and Y.C. helped supervise the study. E.K. supervised the study.

## Notes

The authors declare no potential conflicts of interest.

### Competing Interest Statement

The authors have declared no competing interest.

### Funding Statement

This study was funded by the Department of Defense (HT9425-23-1-0074) and the National Institutes of Health (P50CA211024 and R01CA218668).

### Author Declarations

The Institutional Review Board (IRB) of Weill Cornell Medicine has conducted a review of the study protocol number: 20-06022185, and gave ethical approval for this work

### Summary of Updates

This version of the manuscript has been revised to update the title

